# Reopening Businesses and Risk of COVID-19 Transmission

**DOI:** 10.1101/2020.05.24.20112110

**Authors:** Ashley O’Donoghue, Tenzin Dechen, Whitney Pavlova, Michael Boals, Garba Moussa, Manvi Madan, Aalok Thakkar, Frank J DeFalco, Jennifer P Stevens

## Abstract

**Importance:** The United States has the highest number of confirmed COVID-19 cases in the world, with over 150,000 COVID-19-related deaths as of July 31, 2020^1^. The true risk of a COVID-19 resurgence as states prepare to reopen businesses is unknown.

**Objective:** To quantify the potential risk of COVID-19 transmission in business establishments by building a risk index for each business that measures transmission risk over time.

**Design:** This retrospective case series study uses anonymized cell phone GPS data to analyze trends in traffic patterns to businesses that may be potentially high-risk from January 2020 to June 2020.

**Setting:** Massachusetts, Rhode Island, Connecticut, New Hampshire, Vermont, Maine, New York, and California.

**Participants:** 1,272,260 businesses within 8 states from January 2020 – June 2020.

**Exposure(s):** We monitored business traffic before the pandemic, during the pandemic and after early phases of reopening in 8 states.

**Main Outcome:** Our primary outcome is our business risk index. The index was built using two metrics: visitors per square foot and the average duration of visits. Visitors per square foot account for how densely visitors are packed into businesses. The average duration of visits accounts for the length of time visitors are spending in a business.

**Results:** Potentially risky traffic behaviors at businesses decreased by 30% by April. Since the end of April, the risk index has been increasing as states reopen. On average, it has increased between 10 to 20 percentage points since April and is moving towards pre-pandemic levels of traffic. There are some notable differences in trends across states and industries.

**Conclusion:** Our risk index provides a way for policymakers and hospital decision makers to monitor the potential risk of COVID-19 transmission from businesses based on the frequency and density of visits to businesses. Traffic is slowly moving towards pre-pandemic levels. This can serve as an important metric as states monitor and evaluate their reopening strategies.

## INTRODUCTION

The United States has the highest number of confirmed COVID-19 cases in the world to date, with over 150,000 COVID-19-related deaths as of July 31, 2020^1^. One reason has been the emergence of clusters of COVID-19 from certain events and establishments^2-7^. Monitoring frequency and density of traffic has important implications for policymakers as they decide when and how to safely reopen non-essential businesses^8-9^. A New York Times opinion piece by Baicker et al (2020) found that gyms; full service restaurants and fast food restaurants; and nail salons to have highest visits, longest average length of stay and more opportunities for human-to-human contact before the pandemic, in April 2019, and suggested taking this variability of risk into account when reopening. As businesses reopen with more regulations to ensure social distancing and safety, these 2019 levels of traffic may no longer be a good indicator of reopening risk.

Borg et al, showed an overall average decrease of 25% foot traffic overall in all businesses, with a much smaller drop in religious attendance, during pandemic^10^. Experts have cautioned of the potential resurgence of the virus if we open our economy prematurely^11-15^. However, business traffic patterns after state reopening is still unknown. Further, there may be heterogeneity in traffic patterns by business industry if certain business industries have returned to baseline traffic and operations while others have more regulations in place. The ability to quantify the behaviors during reopening that may be the most prone to increase transmission may help policy-makers make more data-driven decisions when deciding when and how to close and reopen as transmission of COVID-19 fluctuates. Further, many forecasting models use mobility data^17^ to account for social interactions in regions. Often, this mobility data is a broad measure of movement of residents. Our index provides a more granular metric that can quantify human interactions while they are mobile. For example, two regions with the same levels of mobility will likely see very different levels of COVID-19 transmission if one region is practicing social distancing while mobile very seriously and the other is not. Thus, our index can be used in forecasting models to better quantify the social mobility and human interactions in an area, which is an important predictor of transmission and can help to identify a potential second wave.

Because reopening is inevitable, we have constructed a COVID-19 Business Risk Index to monitor the frequency and density of traffic and risk in communities over time as they reopen. This can aid the decision-making process for policymakers and also provide a metric that can be used in retrospectively evaluating various reopening policies.

## RESULTS

Figure 1A displays a map of Maine, New Hampshire, Vermont, Massachusetts, Rhode Island, Connecticut and New York and Figure 1B displays California, with total cumulative COVID 19 cases for each county as of May and locations of potentially high risk business. The darker shades of blue indicate more confirmed COVID-19 cases. The red dots indicate potentially risky business patterns, as defined by a business in top 5% of the index within a state.

**Figure 1A:**
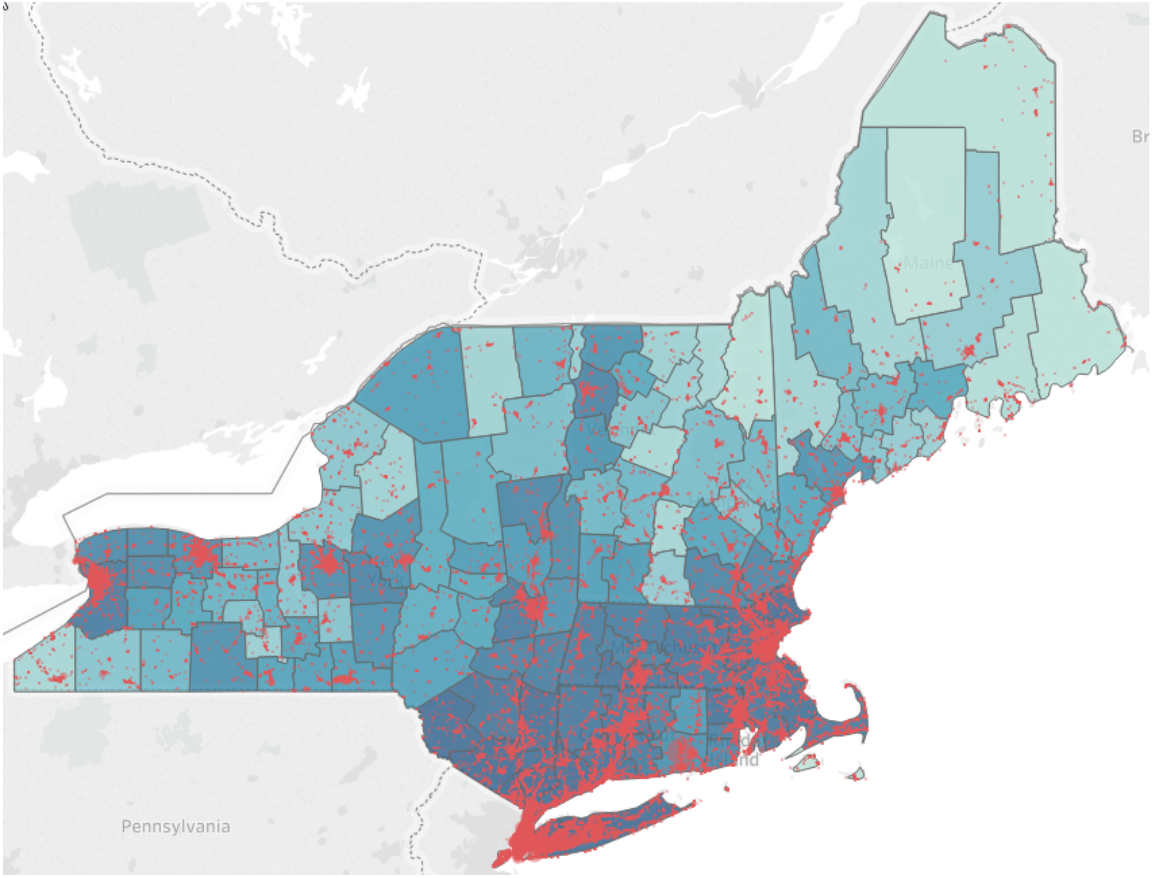
Potential high-risk businesses and COVID-19 cases per 10,000 in the Northeast. The color density of the plots are based on a percentile rank of the total COVID-19 case rates for all counties in the study. Potential high-risk businesses are also displayed on the map as red dots.

**Figure 1B:**
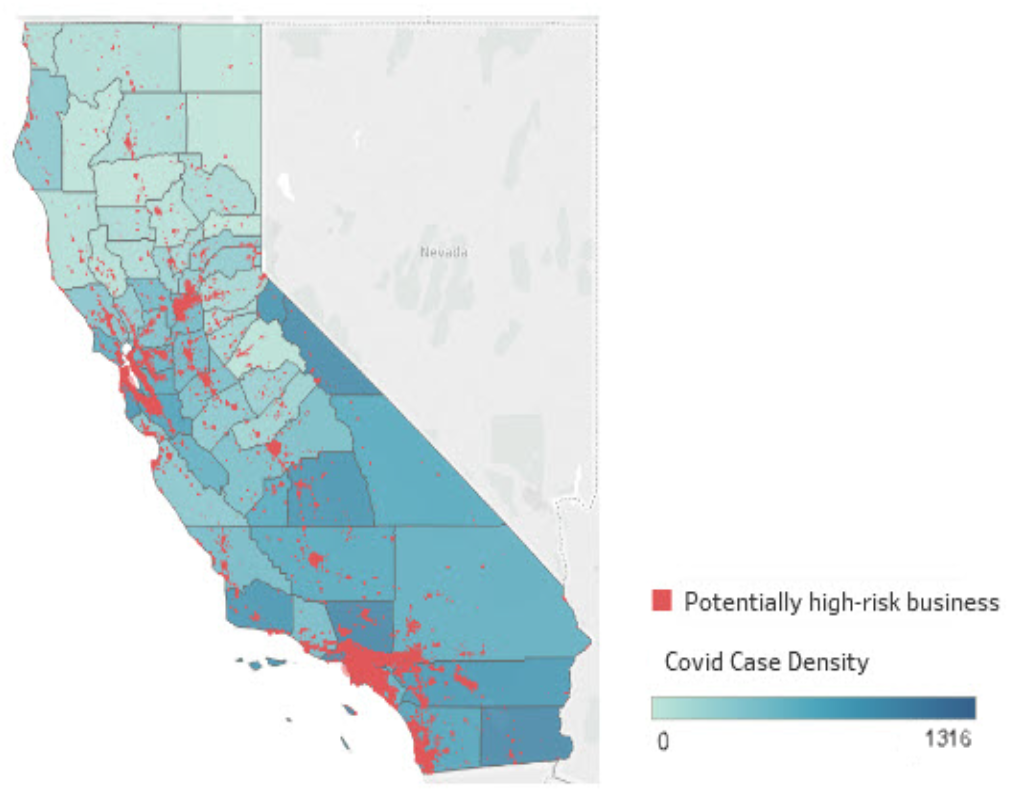
Potential high-risk businesses and COVID-19 cases per 10,000 in California. The color density of the plots are based on a percentile rank of the total COVID-19 case rates for all counties in the study. Potential high risk businesses are also displayed on the map as red dots.

The average business risk in the eight states decreased by 30% (29%-39%) in risky traffic patterns by the beginning of April. In Figure 2, we plot the percent change in the index over time by state. Vermont saw the greatest declines in their index, reaching a low of −40% while the other states in the sample declined by around 30%. Most states have gradually begun increasing these traffic behaviors, and by mid-June many states were operating around −20% (15-22). By mid-June, Maine had returned to −15% of the index.

**Figure 2:**
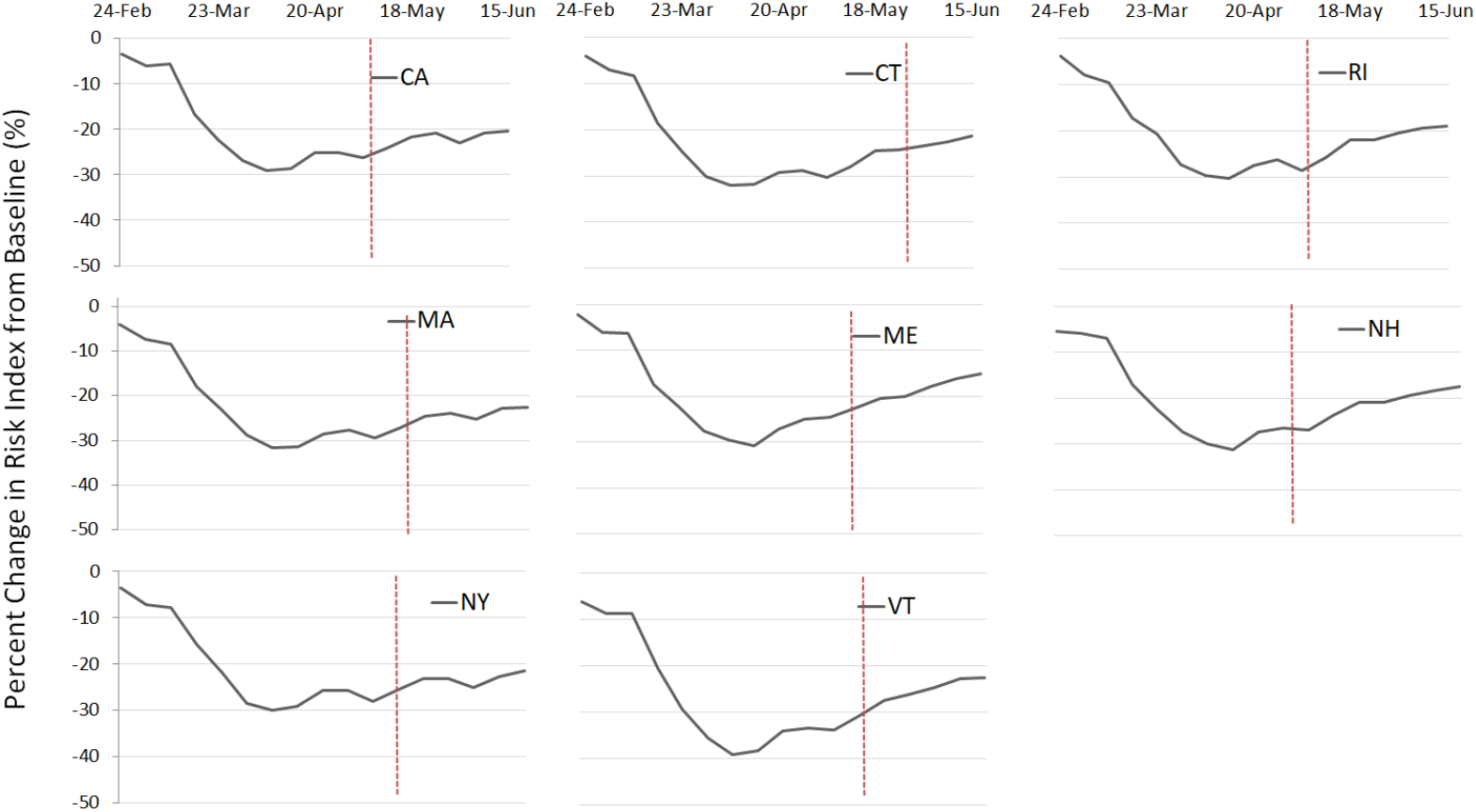
Percent change in risk index by states over time from January 2020 to June 2020. Red line indicates Phase 1 reopening date for each state.

We also looked at risk index over time in four high risk industries; restaurants, bars, universities, and personal care which includes nail and hair salons, barber shops (Figure 3). In all four industries, we see a sharp decrease in risky traffic patterns by the beginning of April. Bars and the personal care industry saw a decrease in average index by 40% whereas restaurants and universities reached a low of −35%. However, since reopening, the risk index has risen for all industries with restaurants only 20% below the pre-pandemic levels by June.

**Figure 3:**
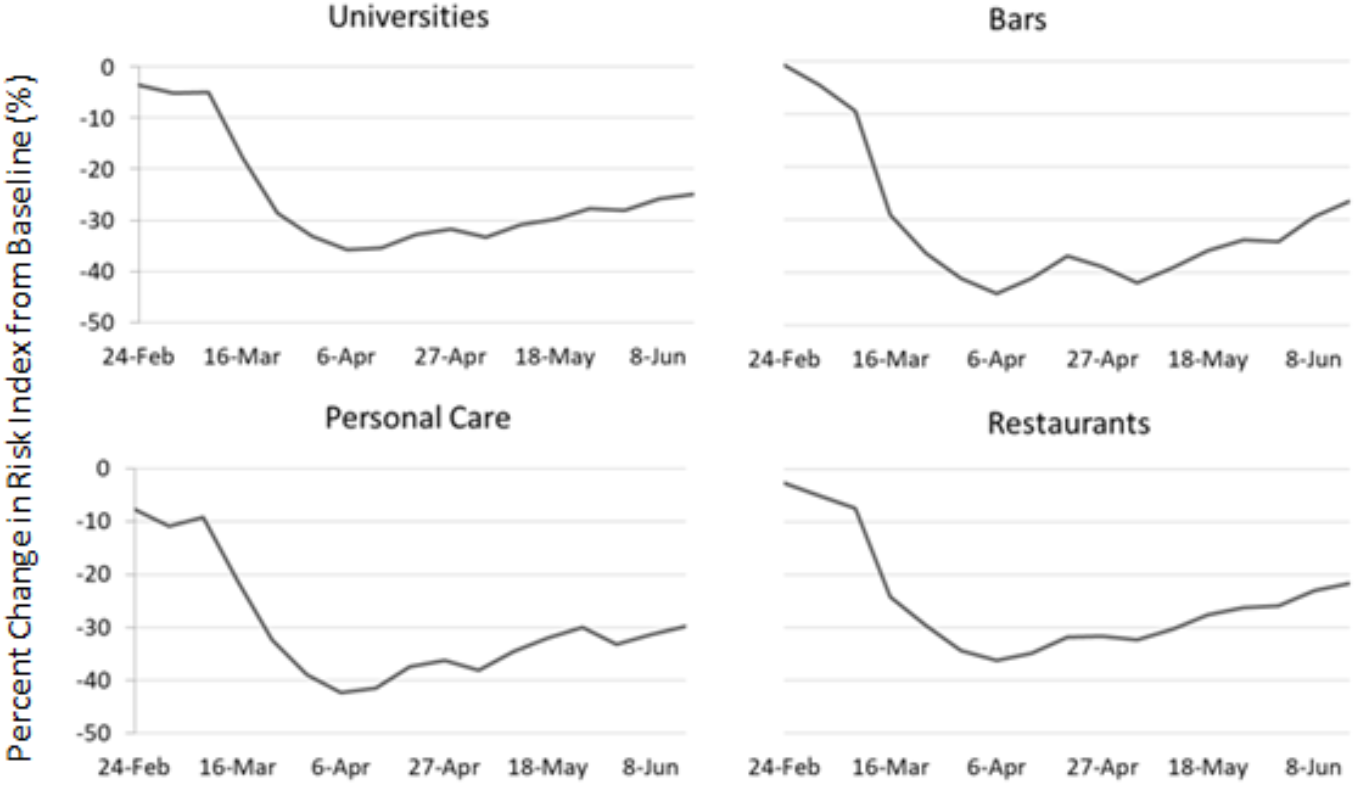
Percent change in index by types of potential high-risk industries over time from Jan 2020 to June 2020.

## DISCUSSION

### HIGH RISK BUSINESSES AND COVID-19

We developed a business risk index to quantify the traffic and risk of COVID-19 transmission at businesses based upon the frequency and duration of visits as well as the density of visitors in the businesses. Businesses with more visitors that stay for longer and are more densely packed are likely to have higher risks of COVID-19 transmission.

Increasing or shifting economic activity across the country may result in more frequent and intense human interactions, which in turn may interact with the prevalence of COVID-19 in a community. While business traffic pre-pandemic and during state-wide shut downs has been studied, business traffic in the “new normal” of reopening is unknown. We propose that tracking how individuals use different businesses may inform policymakers to reopen different businesses the safest way possible. This study can also be useful for hospital decision makers. Monitoring traffic at businesses in their service area and to these businesses may help hospitals prepare for a potential second-wave if the risk of business traffic is high. For example, metrics for mobility in a region have been incorporated into many forecasting models. Often, these metrics measure mobility broadly and don’t include the levels of potential human interaction. It’s important to quantify mobility through the lens of human interaction to take into account, for example, that mobility to an outdoor park where everyone is socially distant will be different than mobility to a crowded, indoor business. Our index can provide prediction models an improved measure of social mobility for forecasting potential second waves.

### LIMITATIONS

There are several limitations to this study. Because Safegraph’s location data uses GPS location data, there is room for error. Businesses that occupy one floor of a multi-level building may have artificially high visits per square feet because GPS cannot differentiate between floors. Locations that are very small and closely located next to other businesses may erroneously have spillovers of their visits to nearby stores or visits from other nearby stores attributed to them, if the GPS does not pinpoint them precisely. SafeGraph’s data accounts for individuals who have a cell phone with location services. Finally, while traffic moves towards pre-pandemic levels as states reopen, states may experience different effects of reopening based upon other practices such as mask adherence and fiberglass barriers at businesses that did not exist pre-pandemic.

### FUTURE WORK

We are building an online decision-support tool that will allow policymakers and hospital decision makers to visualize potential high risk businesses in their area and monitor weekly risk to these businesses. We are currently testing a prototype of our tool at a health system in New York to monitor business traffic (e-figures 1-4) and our index is being integrated into a forecasting model for a large health system in Massachusetts to help predict a potential second wave. Our tool is helping policymakers and hospital decision makers monitor the risks in their community as they reopen.

Finally, as states reopen non-essential businesses in phases, we plan to evaluate the effects of various reopening policies on COVID-19 transmission using our risk index. Knowing the effects of reopening can help future policymakers and hospital decision makers plan for the potential impact of reopenings and quantify the effectiveness of various reopening policies in a data-driven way.

## METHODS

### DATA AND SETTING

We used data from SafeGraph Monthly Patterns from January 2020 – June 2020 and SafeGraph Core Points of Interest data to measure business characteristics and traffic. SafeGraph is a data company that aggregates anonymized location data from numerous applications in order to provide insights about physical places. To enhance privacy, SafeGraph excludes census block group information if fewer than five devices visited an establishment in a month from a given census block group. This study focused on counties in 8 states (Massachusetts, Rhode Island, Connecticut, New Hampshire, Vermont, Maine, New York, and California). We examine traffic to 1,272,260 businesses from January 2020 – June 2020.

### STUDY VARIABLES

We extracted weekly measures of number of visits, square feet of businesses and bucket dwell times. Bucketed dwell times report the number of visitors that week whose visit fell within several buckets of time.

### INDEX CONSTRUCTION

We calculated a baseline measure of the index for each business in our sample prepandemic in order to quantify what “normal” traffic looks like at these locations. We monitored the risk index weekly from January 2020 - June 2020 to track the riskiness of communities as they shut down and then reopen.

The index was built using two metrics: visitors per square foot and the average duration of visits. Visitors per square foot account for how densely visitors are packed into businesses. Businesses that are more densely packed may have a higher risk of COVID-19 transmission. The average duration of visits accounts for the length of time visitors are spending in a business. Businesses where visitors linger for longer periods of time could be riskier for COVID-19 transmission than businesses where visitors are quickly in and out of the business^8, 10^.

Visitors per square foot is calculated by dividing the number of visitors by the square footage of the business. The average duration of visits is calculated by first taking the centroid of each bucket of dwell time and multiplying by the number of visitors in that bucket to estimate the total dwell time at that business across all visitors. Then, this total dwell time is divided by the number of visits to generate the average duration of a visit in minutes.

Our composite risk index incorporates these two metrics and is normalized to fall between 0 and 100. Because our data comes from cell phone GPS location data, there are some limitations, which we describe in more detail in the limitations section, that contribute to large outliers. For this reason, we exclude outliers whose visits per square feet and average duration of visits fall more than 8 times the interquartile range above the third quartile. This excludes 31,909 observations (1.8% of the total observations).

We normalize both metrics by state to fall between 0 and 100 by subtracting the minimum value and then dividing by the range and multiplying by 100. We calculate these by state so that the index is not clouded by differences in traffic patterns across states. For example, we don’t want to compare businesses in New York directly to businesses in Maine. Normalizing the metrics for each state allows us to only compare businesses within the same state.

Next, we sum these normalized metrics for visitors per square foot and average duration of visits. This sum is then normalized by state to fall between 0 and 100 by subtracting the minimum value and then dividing by the range and multiplying by 100. The resulting value between 0 and 100 is our composite COVID-19 Business Risk Index.

### STATISTICAL ANALYSIS

Statistical analysis was performed using Stata SE version 14.2 (StataCorp) and SAS (v. 9.4, SAS Institute Inc., Cary, NC). We examined changes in the index over time by state. We also stratified by industry to explore which industries had the most potentially high-risk businesses.

## Data Availability

The Johns Hopkins University COVID-19 data and the New York Times COVID-19 data are publically-available, linked below. SafeGraph's Monthly Patterns data and Core POI data are restricted access. The documentation for SafeGraph's datasets and the link to apply for access are linked below.

https://github.com/CSSEGISandData/COVID-19

https://github.com/nytimes/covid-19-data

https://docs.safegraph.com/docs/places-schema#section-patterns

https://docs.safegraph.com/docs#section-core-places

https://www.safegraph.com/weekly-foot-traffic-patterns

## DATA AVAILABILITY

The data that support the findings of this study are available from SafeGraph but restrictions apply to the availability of these data, which were used under license for the current study, and so are not publicly available. Data are however available from the authors upon reasonable request and with permission of SafeGraph.

## SUPPLEMENTAL FIGURES

**e-Figure 1:**
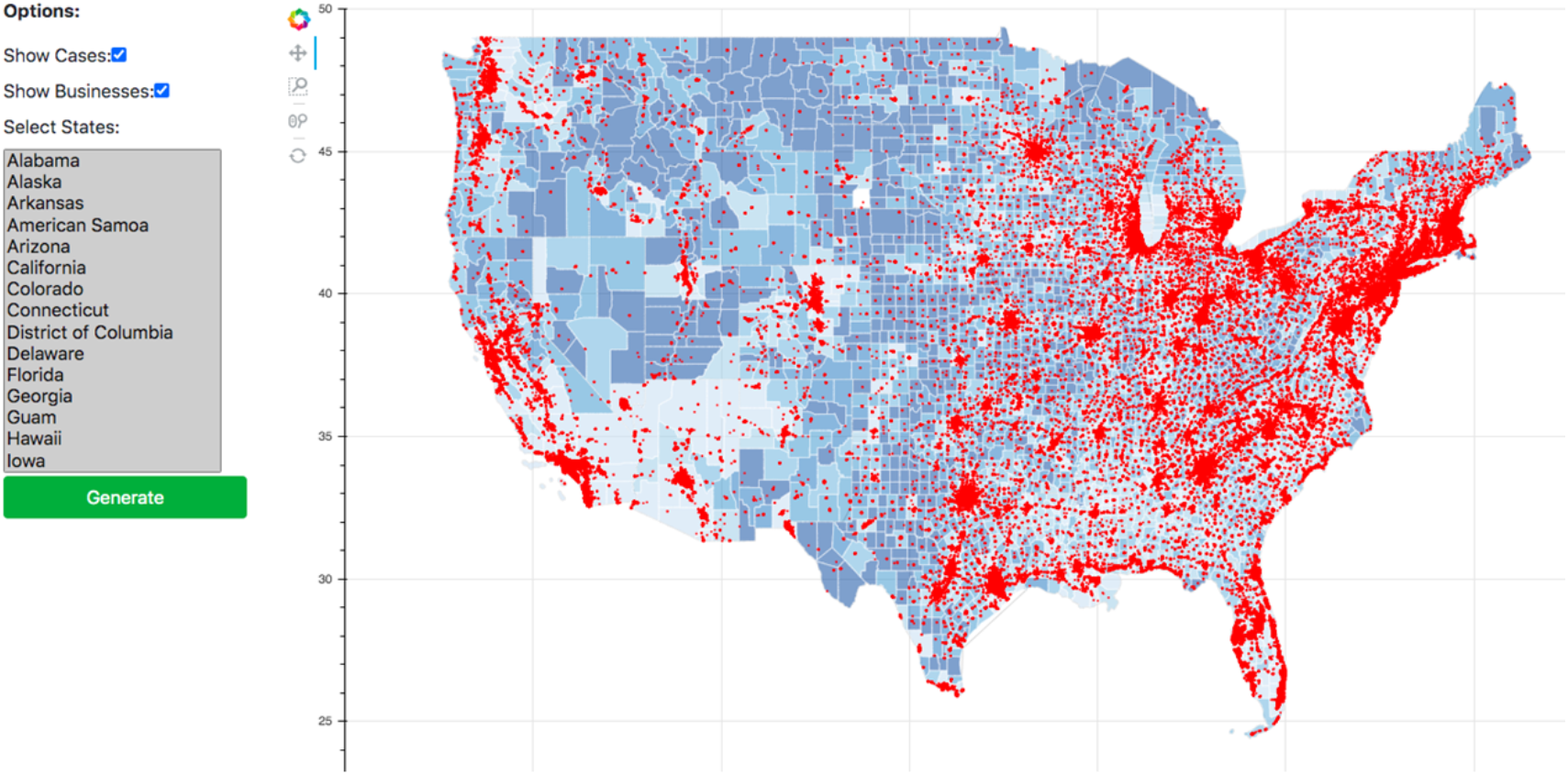
Prototype of main page of interactive dashboard for all states. Darker shades of blue indicates more covid cases. Red indicates a potentially high risk business location. The red dots can be filtered by risk level, percentage change in risk over time, or by industry. This view displays states by the business risk index and covid cases by county. One can filter on state as in e-Figure 2.

**e-Figure 2:**
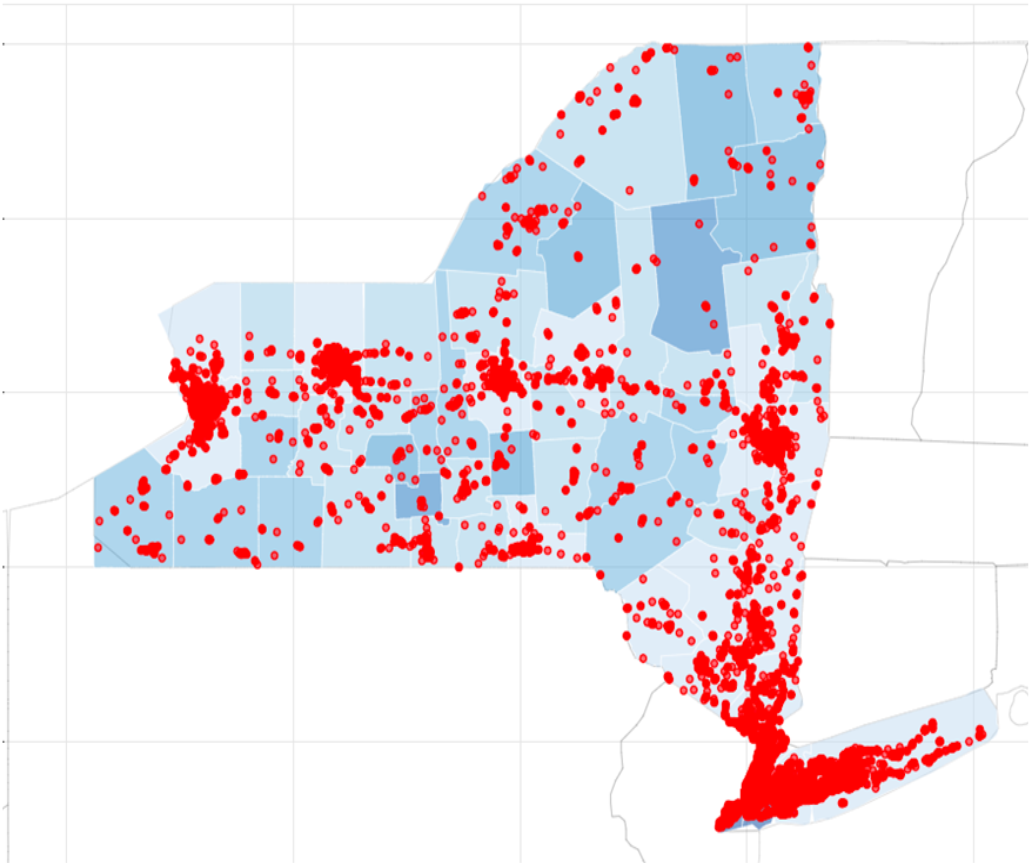
Prototype of main page of interactive dashboard for New York. Darker shades of blue indicates more covid cases. Red indicates a potentially high risk business location. The red dots can be filtered by risk level, percentage change in risk over time, or by industry. This view displays the dashboard filtered to show a map of New York.

**e-Figure 3:**
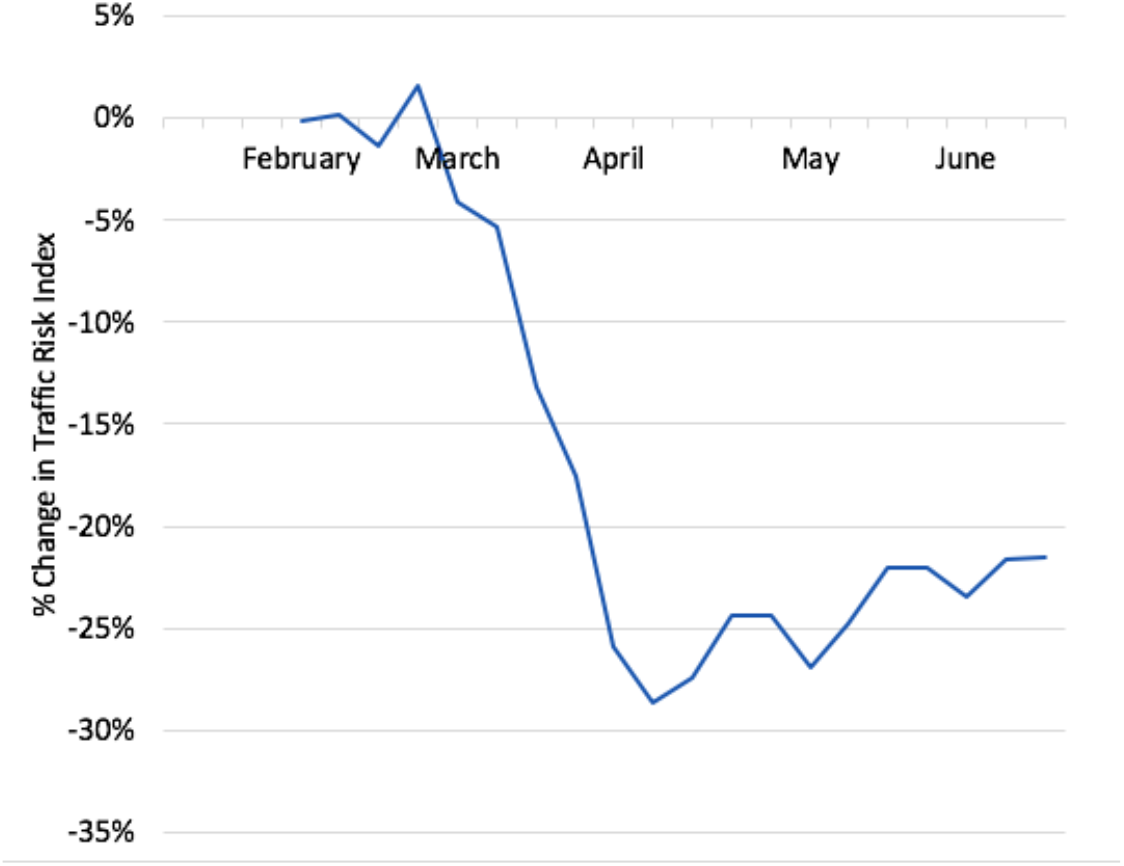
Detailed view of dashboard of percentage change in business risk index over time by census tract. After filtering by state, business risk can be displayed in several different ways. This view displays change in business risk index over time by county or census tract.

